# Assessing the feasibility of Nipah vaccine efficacy trials based on previous outbreaks in Bangladesh

**DOI:** 10.1101/2020.12.06.20244871

**Authors:** Birgit Nikolay, Marc Lipsitch, Mahmudur Rahman, Stephen P. Luby, Henrik Salje, Emily S. Gurley, Simon Cauchemez

**Affiliations:** Mathematical Modelling of Infectious Diseases Unit, Institut Pasteur, UMR2000, CNRS, 75015 Paris, France; Harvard T.H. Chan School of Public Health, Boston, Massachusetts, USA; Consultant; Infectious Diseases and Geographic Medicine Division, Stanford University, Stanford, California, USA; Cambridge Infectious Diseases, University of Cambridge, Cambridge, UK; Department of Epidemiology, Johns Hopkins Bloomberg School of Public Health, Baltimore, Maryland, USA

**Keywords:** Nipah virus, vaccine trial, emerging pathogens

## Abstract

**Background:** Nipah virus (NiV) is an emerging, bat-borne pathogen that can be transmitted from person-to-person. Vaccines are currently being developed for NiV, and studies funded to evaluate their safety and immunogenicity, so that they could possibly be used to contain outbreaks. An important unanswered question is whether it will be possible to evaluate the efficacy of vaccine candidates in phase III clinical trials in a context where spillovers from the zoonotic reservoir are infrequent and associated with small outbreaks. The objective of this study was to investigate the feasibility of conducting a phase III vaccine trial in Bangladesh, the only country reporting regularly NiV cases.

**Methods:** We used simulations based on previously observed NiV cases from Bangladesh, an assumed vaccine efficacy of 90% and other NiV vaccine target characteristics, to compare three vaccination study designs: (i) cluster randomized ring vaccination, (ii) cluster randomized mass vaccination, and (iii) an observational case-control study design.

**Results:** The simulations showed that, assuming a ramp-up period of 10 days and a mean hospitalization delay of 4 days, it would take 516 years and over 163,000 vaccine doses to run a ring vaccination trial under current epidemic conditions. A cluster-randomized trial in the two most affected districts would take 81 years and 2.3 million vaccine doses. An observational case-control design in these two districts would require seven years and 2.5 million vaccine doses.

**Discussion:** Without a change in the epidemiology of NiV, ring vaccination or cluster-randomized trials are unlikely to be completed within a reasonable time window. In this light, the remaining options are: (i) not conducting a phase III trial until the epidemiology of NiV changes, (ii) identifying alternative ways to licensure such as observational studies or controlled studies in animals such as in the US Food and Drug Administration’s (FDA) Animal Rule.

## Introduction

Vaccines can contribute to controlling the spread of emerging pathogens, but the development of such vaccines is hampered by their limited commercial value and an unclear path to licensure due to difficulties in designing trials in the context of small or unpredictable case numbers. For example, at the start of the Ebola outbreak in West Africa, several candidates were promising in animal studies but none had been given to humans because of the small size and unpredictability of prior outbreaks. This resulted in lengthy delays in being able to use the vaccine for outbreak control [1]. These difficulties were one motivation for the creation of the Coalition for Epidemic Preparedness Innovations (CEPI), with the mission to accelerate the development of vaccines through proof-of-concept, safety, and immunogenicity studies, to enable efficient efficacy testing and licencing in the case of a health emergency, and initiatives to develop vaccines for emerging infectious diseases where there was no clear pathway to a phase III clinical trial [2]. CEPI has announced financing for the development of vaccine candidates against seven emerging pathogens prioritized by the World Health Organization (WHO) including severe acute respiratory syndrome coronavirus 2, Ebola virus, Middle East respiratory syndrome coronavirus (MERS-CoV), Lassa virus, Rift Valley fever virus, chikungunya virus, and Nipah virus (NiV), as well as for capacity building to develop vaccines against an unknown disease “X” [3,4].

NiV is an emerging, bat-borne pathogen that can also be transmitted from person-to-person [5,6]. While studies are currently funded to evaluate the safety and immunogenicity of four NiV vaccine candidates [4], an important question is whether, under current conditions, vaccine candidates shown to be safe and immunogenic could be tested for efficacy in phase III trials or whether alternative pathways to licensure are needed. The question arises because humans acquire NiV infections from the zoonotic reservoir infrequently and these infections are associated with outbreaks that are small and often detected late [6]. Bangladesh appears to be the most suitable place for such trials, as the epidemiology there is relatively well understood, and it is the country that reports the most NiV outbreaks [6]. However, even in Bangladesh, NiV cases remain rare: the country reports seven spillovers from the zoonotic reservoir into human populations per year on average [7]; outbreaks have never extended beyond five generations or 34 cases; and only ∼10% of cases transmit the virus to another person [6].

Computational simulations of disease outbreaks can inform the design of vaccine trials and have been previously used to identify the optimal trial strategy during the Ebola outbreak in West Africa and for other aspects of vaccine trial design and interpretation [8–10]. To draw valid conclusions, it is important that these simulations accurately reproduce key outbreak characteristics, which can be difficult for emerging pathogens where detailed knowledge about transmission dynamics and natural history of infection is unavailable [11]. For NiV, such information has been collected for more than 10 years of routine surveillance in Bangladesh [6] and can now be used to inform simulations that reproduce NiV transmission and allow to test different trial designs. The aim of this study was to assess the feasibility of a phase III vaccine trial for NiV considering the current epidemiological characteristics of the pathogen. We compared three study designs: (i) cluster randomized ring vaccination, (ii) cluster randomized mass vaccination, and (iii) an observational case-control study design.

## Methods

### Simulated trial designs

We simulated zoonotic cases (i.e., humans who acquired infections from the bat reservoir) and secondary human cases and three phase-three vaccine design approaches (Figure 1).

**Figure 1.**
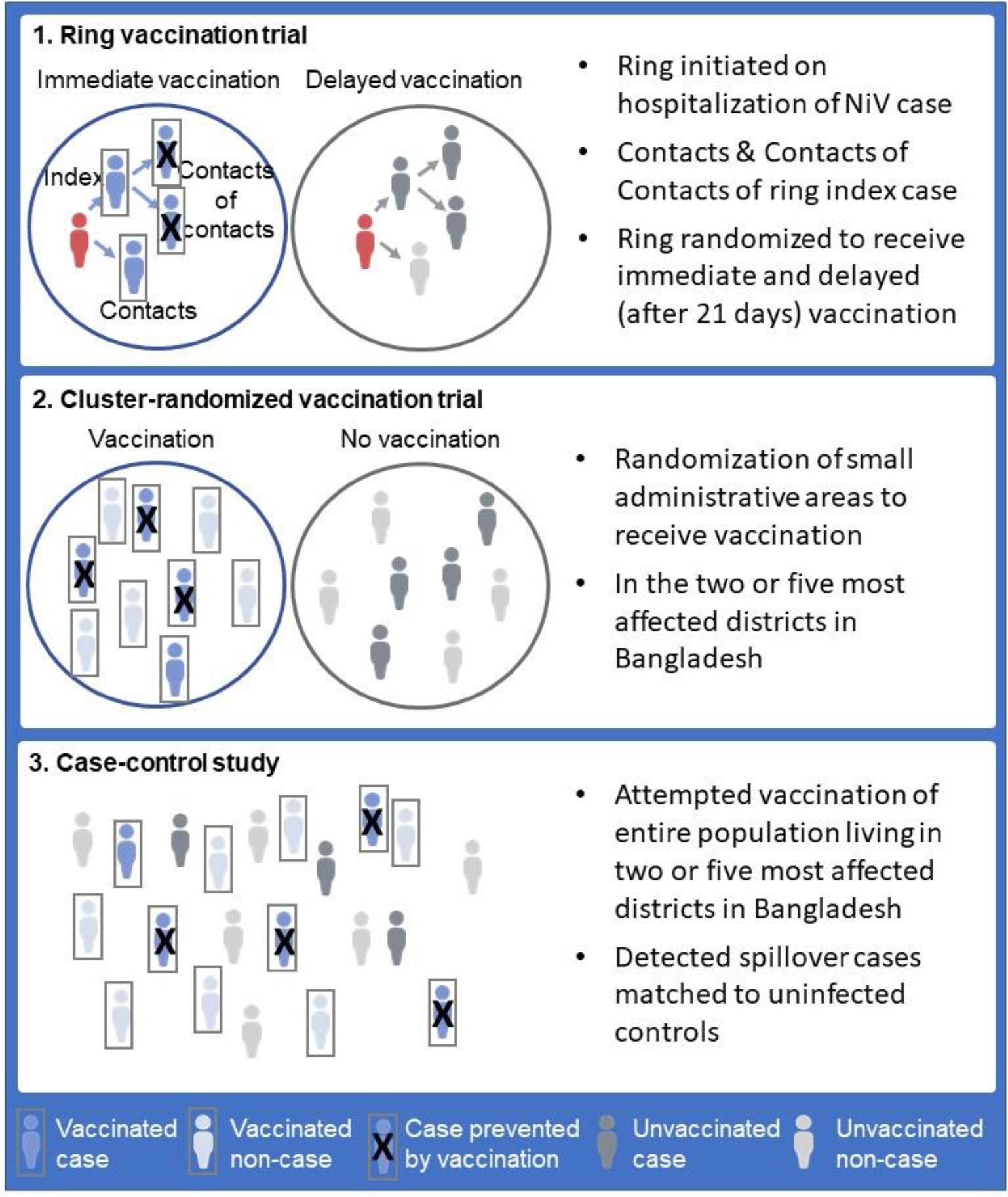
Considered study designs.

In the simulated ring vaccination trial, the hospitalization of any NiV case resulted in the formation of a ring, defined as contacts and contacts of contacts of the ring’s index case-patient. The hospitalization of an infected contact of a contact resulted in the initiation of a new ring. Rings were randomized to receive immediate or delayed (21 days) vaccination and the number of detected cases within a pre-specified time window was compared between the study arms to estimate the vaccine efficacy. This design is broadly similar to that of the Ebola ça Suffit! trial used to test an Ebola virus disease vaccine in Guinea in 2014-5 [12].

In the cluster randomized mass vaccination trial design, individuals were randomized at the level of small administrative areas to receive vaccination. Two geographic scales for enrollment were considered: (i) the two districts (Faridpur and Rajbari districts, with a total population of ∼3 million individuals), or (ii) the five districts (additionally including Naogaon, Rangpur, and Gopalganj districts, with a total population of ∼6.8 million individuals) with highest reported detection rates of spillovers from the zoonotic reservoir into humans [7]. The vaccine efficacy was estimated by comparing the total number of cases detected by study arm.

In the case-control study design, vaccination was offered to the entire population living in the selected districts (the same geographic areas are considered as for the cluster randomized trial design). Detected zoonotic cases in these districts were then matched to uninfected controls and the vaccination status among enrolled cases and controls was used to estimate the vaccine efficacy (1-odds ratio of vaccination among cases compared to controls) [13].

### Vaccine characteristics

Vaccine efficacy is the probability that a zoonotic or secondary case is prevented because that individual has been vaccinated. As characteristics of future Nipah vaccine candidates are unknown we based the assumptions on vaccine efficacy on the Nipah vaccine target product profile defined by the World Health Organization [WHO]; i.e. a vaccine efficacy of 90% (preferred target product profile) or 70% (minimal target product profile) [14]. We further assumed that vaccine efficacy reaches its maximum after a single dose and a ramp-up period (5 to 15 days) during which efficacy is linearly increasing and that no waning of vaccine-induced immunity occurs. We also considered vaccine post-exposure effects, i.e., prevention of symptoms and transmission if vaccination occurs within 1-5 days of exposure, as previously observed in animal studies of a vesicular stomatitis virus-based NiV vaccine candidate and in many licensed viral vaccines [15,16]. For population-based studies (cluster randomized trials and case control studies), we assumed that vaccination is rolled-out every 1-10 years (vaccination frequency) resetting the vaccination coverage to 70% or 90%. We allowed the vaccination coverage to vary over time dependent on births of susceptible individuals and deaths of vaccinated or susceptible individuals.

### Simulating NiV zoonotic cases and interhuman transmission chains

We simulated NiV zoonotic cases in Bangladesh assuming a Poisson distribution with a mean annual zoonotic spillover detection rate of 0.04 per 1 million people as reported in 2007-2018 [7]. We further simulated zoonotic cases in the two districts (average annual zoonotic spillover detection rate of 0.7 per 1 million people) or the five districts (average annual zoonotic spillover detection rate of 0.1 per 1 million people) with highest reported zoonotic spillover detection rates [7]. We accounted for population dynamics assuming predicted annual birth and death rates for Bangladesh [17].

We simulated interhuman transmission of NiV following the occurrence of a zoonotic case until the end of the transmission chain using a branching process model. For each NiV case we drew the number of secondary cases (i.e. cases infected through interhuman transmission) from a negative binomial distribution with reproduction number R=0.20 and overdispersion parameter k=0.06 as observed for cases reported during 2007-2014 (a time period of systematic NiV surveillance) [6,7]. For each secondary NiV case, we drew the time from disease onset in the infector to infection from a discretized gamma distribution with mean 4 days and standard deviation (SD) 2 days and the time from infection to symptom onset in the secondary case from a gamma distribution with mean 10 days and SD 2 days [6]. We drew the time from disease onset to hospitalization of a case from a discretized gamma distribution with mean 5 days and SD 2 days (as reported in Bangladesh during 2001-2014 [6]) or with mean 1 day and SD 1 day (a scenario of minimal case detection delay), and assumed that case hospitalization had no impact on interhuman transmission [6].

### Simulating vaccination studies

For the ring vaccination trial, we simulated 50,000 rings per trial arm to estimate the intraclass correlation coefficient (ICC) assuming a ring size of 50 individuals in the baseline scenario [18]. We then estimated the required number of rings for a study powered with 80% (and a level of significance alpha=0.05) based on the cumulative number of cases occurring in each arm within a 15-36 day window following vaccination (starting at the average incubation period plus the ramp-up period and ending 21 days later), allowing the ICC to vary between study arms [18]. We estimated the study duration based on the median time to observe the required number of rings in 1,000 simulated studies. We simulated studies for up to 150 years; if the number of required rings was not reached, we estimated the duration based on the average number of rings enrolled per year in the simulations.

For the cluster randomized controlled trial, we simulated 50,000 clusters per trial arm for different trial durations (10-200 years) to estimate the ICC, assuming a cluster size of 100,000 individuals in the baseline scenario and that transmission events only occur within a given cluster. We quantified the required number of clusters based on the cumulative number of cases occurring in each arm [18]. We then used regression analysis to identify the trial duration that results in a total sample size corresponding to the population of selected districts (Supplementary material).

For the case-control study, we estimated the number of zoonotic cases required to perform a case-control study powered with 80% (and a level of significance alpha=0.05; one-sided test) using methods implemented in the epiR package and described by Dupont [19]. As vaccination coverage varied over time, we used the observed vaccination coverage resulting in the largest sample size. We then estimated the study duration based on the median time to observe the required number of cases in 1,000 simulated studies.

All parameter values for baseline scenarios and sensitivity analyses and their sources are summarized in Table S1 in the Supplementary material. We also assessed how improvements in case detection may reduce study duration [20].

## Results

### Cluster randomized controlled ring vaccination trial design

Assuming a ramp-up period of 10 days and a mean hospitalization delay of 4 days, 1,807 rings per study arm need to be enrolled to run the trial for a 90% efficacious vaccine (Figure S1), which takes around 516 years and over 163,000 vaccine doses (Figure 2 A). Even in a more optimistic scenario, where NiV cases are hospitalized on average 1 day after disease onset, the study has to run approximately 317 years and requires at least 100,000 doses. There are further reductions in study duration if the vaccine has significant post-exposure effects and a shorter ramp-up period. However, even for a vaccine with 5 days post-exposure effects and only 5 days ramp-up period, the study duration remains 110 years and requires 35,000 doses (Figure 2 A). In the latter scenario, the proportion of contacts whose infection is prevented increases from 7% (no post-exposure and 10 day ramp-up) to 62% (Figure S2); infections among contacts of contacts, which without post-exposure effects are preventable at a percentage close to the vaccine efficacy, are only rarely observed for NiV (Figure S2). Study duration and number of doses for a 70% efficient vaccine are shown in Figure 2 B.

**Figure 2.**
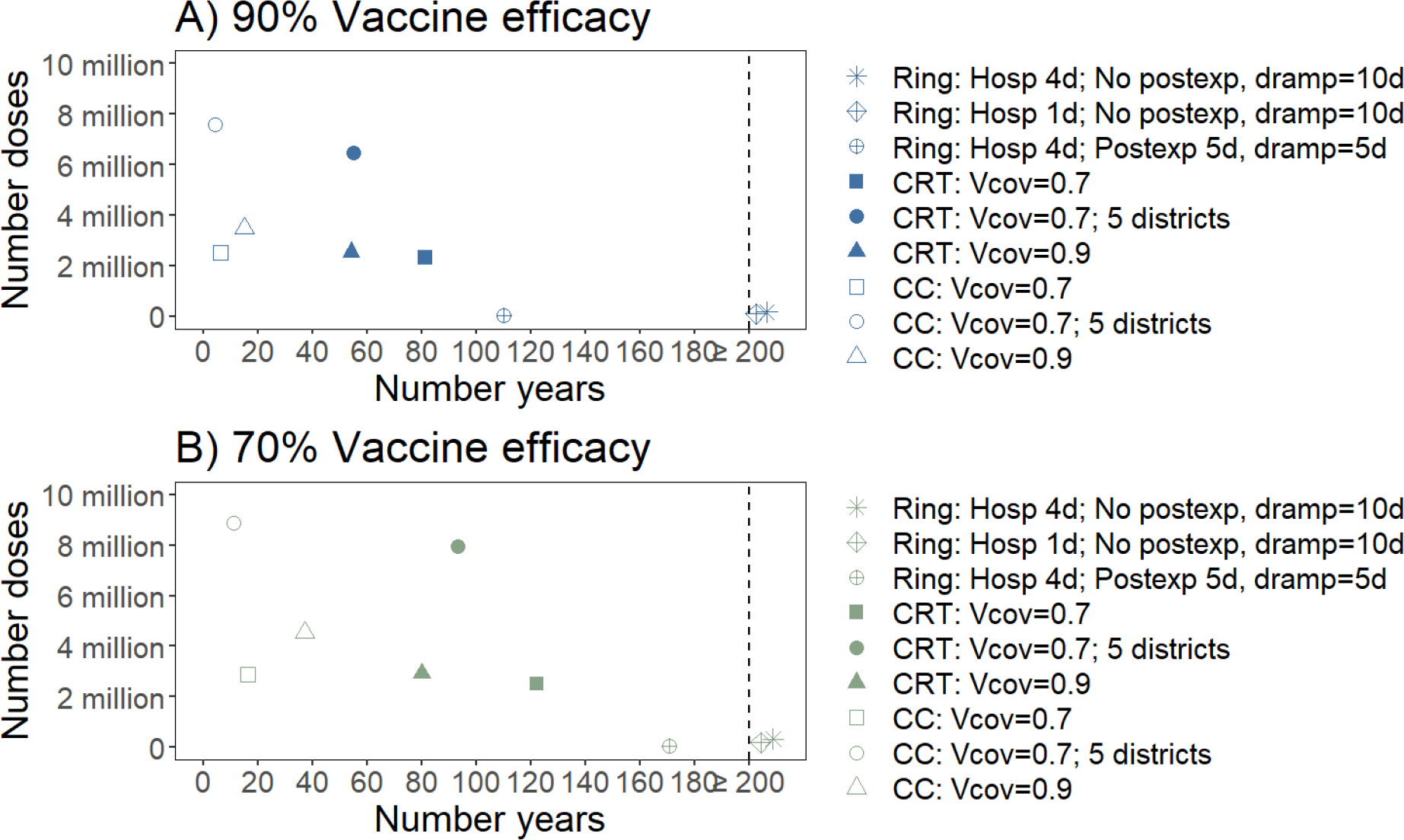
Number of years and vaccine doses required for different study designs assuming a vaccine efficacy of 90% (A) and 70% (B). Estimates are presented for a ring size of 90 individuals (ring vaccination trial), a cluster size of 100,000 individuals and a vaccination frequency of five years (cluster randomized trial and case control study), and ten controls per case (case control study). Ring vaccination trial (Ring); Cluster Randomized Trial (CRT); Case control study (CC); Mean hospitalization delay (Hosp); Post-exposure duration (Postexp); Vaccine coverage (Vcov); Duration of ramp-up period (dramp). Study durations are truncated at 200 years.

### Cluster randomized controlled trial design

With a vaccine efficacy of 90% and a vaccination coverage of 70% in the vaccination arm, a cluster randomized trial involving the total population of Faridpur and Rajbari districts (15 clusters per trial arm) requires 81 years and 2.3 million vaccine doses (Figure 2 A). To reduce the study duration to 10 years, 163 clusters per arm would need to be enrolled, representing 10 times the population of the two districts (Figure S3). If we extend the study area to three additional districts, the study requires 55 years and 6.5 million vaccine doses (Figure 2 A). A vaccination coverage of 90% results in a study duration of 54 years and 2.5 million vaccine doses (Figure 2 A). If the vaccine is 70% efficacious, study durations are substantially longer (e.g. 122 years for a vaccination coverage of 70% in Faridpur and Rajbari districts) (Figure 2 B).

### Case-control study design

For a vaccine efficacy of 90% and a vaccination coverage of 70%, a case-control study with ten controls per case requires six NiV zoonotic cases (Figure S4). If implemented in Faridpur and Rajbari districts, the study takes around seven years and 2.5 million vaccine doses, compared to five years and 7.6 million vaccine doses if the study area is extended to three additional districts (Figure 2 A). For a vaccination coverage of 90%, nine NiV zoonotic cases need to be observed, which takes 17 years and 3.7 million vaccine doses. If the vaccine is 70% efficacious, the study requires 17 years and 2.9 million vaccine doses (70% vaccination coverage) (Figure 2 B).

### Improving case detection

It has been estimated that the true number of NiV cases in Bangladesh may be double that observed [20]. Even if the number of detected zoonotic cases doubled, a ring vaccination trial for a 90% efficacious vaccine (10 days ramp-up, no post-exposure effects) would require 258 years (Figure 3). A cluster randomized trial in Faridpur and Rajbari districts with a vaccine coverage of 70% would require 52 years and a case control study three years (Figure 3).

**Figure 3.**
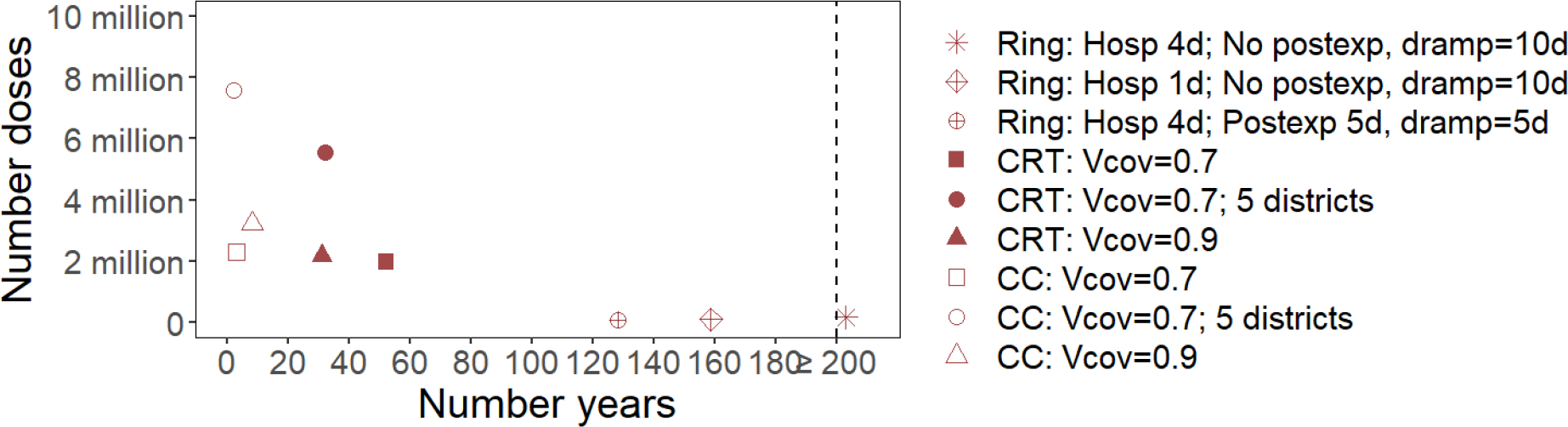
Number of years and vaccine doses required for a 90% efficacious vaccine if the number of detected spillovers from the zoonotic reservoir into human populations doubled. Estimates are presented for a ring size of 90 individuals (ring vaccination trial), a cluster size of 100,000 individuals and a vaccination frequency of five years (cluster randomized trial and case control study), and ten controls per case (case control study). Ring vaccination trial (Ring); Cluster Randomized Trial (CRT); Case control study (CC); Mean hospitalization delay (Hosp); Post-exposure duration (Postexp); Vaccine coverage (Vcov); Duration of ramp-up period (dramp). Study durations are truncated at 200 years.

Conducting a vaccine efficacy study within a window of 10 years, for a ring vaccination trial, would only be feasible if more than 50 times as many zoonotic cases were detected per year, which is substantially in excess of the total number of underlying zoonotic cases estimated to occur [20]. For a cluster randomized trial in Faridpur and Rajbari districts, eight times as many zoonotic cases would need to be detected per year.

### Sensitivity analysis-effect of parameter assumptions on study duration

For the ring vaccination design, increasing the size of rings has no effect on study duration (Figure S5). A longer duration of vaccine ramp-up (e.g. 327 years for 5 days vs. 1025 years for 15 days) (Figure S5) and an earlier peak in case infectivity result in longer study duration (e.g. 516 years for a mean of 4 days vs. 794 years for a mean of 2 days) (Figure S6). For the cluster randomized design, increasing cluster sizes has no effect on study duration while a lower vaccination frequency results in slightly longer study duration (e.g. 81 years for vaccination every 5 years vs. 88 years for vaccination every 10 years) (Figure S7). For the case-control design, increasing the number of enrolled controls per case to more than ten (e.g. 6 years for 10 controls or 15 controls) and increasing the vaccination frequency has little effect on study duration (e.g. 6 years for vaccination every 5 or 10 years) (Figure S8).

## Discussion

Our findings suggest that, in Bangladesh, ring vaccination or cluster-randomized trials are unlikely to be completed within a reasonable time window with current epidemiologic characteristics. The ring vaccination trial design, which has been a successful strategy for the evaluation of Ebola virus vaccine candidates, is unsuitable for NiV under current conditions, due to short interhuman transmission chains. In this light the global community has the following options to consider: (i) not conducting a phase III trial until the epidemiology of NiV changes, potentially resulting again in major delays in the use of the vaccine if a larger outbreak occurs, (ii) identifying alternative ways to licensure of the vaccine not involving clinical trials in humans through either observational studies or controlled animal studies as in the US Food and Drug Administration’s (FDA) Animal Rule.

An observational case-control design may represent a viable alternative strategy, which we estimate could be completed within 7 years, even with no investments to improve case detection. Observational case-control trials are complicated by concerns about potentially biased vaccine efficacy estimates as vaccinated and unvaccinated individuals may differ in other risk factors [21]. Such biases can however be minimized by considerations in the study design e.g. through matching of controls to cases in key characteristics, such as sociodemographic characteristics, or other risk factors including occupational exposure or geographic location. A “test-negative” design, where laboratory confirmed NiV patients are considered as cases and suspected NiV patients that tested negative are considered as controls, could help preventing some biases related to healthcare seeking behavior [21–23]. Evaluating vaccine efficacy based on an alternative outcome that cannot be causally affected by the vaccine can help testing for potential biases in the study [21]. Case-control designs have been previously used to evaluate vaccine efficacy in the context of routine vaccination programs, as in the case of tuberculosis, meningococcus, measles and poliomyelitis vaccines [24–28]. A practical advantage of the case-control study design is that it may raise fewer ethical concerns for participants, as vaccination is not withheld from a part of the population that would have been vaccinated otherwise, and could therefore be more acceptable to communities. A case-control design may also be used to evaluate vaccine safety by including an active or passive surveillance component for adverse vaccine effects [24]. Such observational designs can however only be attempted after the vaccine is made available for mass vaccination through some regulatory process (likely short of licensure given a lack of efficacy data).

Currently, four NiV vaccine candidates are being funded by CEPI to undergo safety and immunogenicity testing with the idea that safe and immunogenic vaccines could be used – and tested – in the context of any future, large outbreaks. Under current epidemiologic scenarios, simultaneous testing of more than one vaccine candidate in phase III trials would lead to substantially longer study durations. Considering the practical limitations highlighted here, it may therefore be necessary to focus efforts to evaluate efficacy of just one vaccine candidate, and criteria for deciding on which candidate to pursue would need to be developed.

Model assumptions used to simulate zoonotic NiV cases and interhuman transmission influenced the estimated study durations; wherever possible, our assumptions are made using data and we consistently used optimistic scenarios from the perspective of our ability to conduct a trial within a reasonable timeframe. For example, in ring vaccination trials, we assumed that hospitalization of cases had no impact on NiV transmission, which is justified as infection control measures in Bangladeshi hospitals are often limited [29]. In some settings, hospitalization may lead to increased transmission due to superspreading events as observed in two hospital based outbreaks in India [30,31]. Further, we chose an optimistic scenario where NiV cases are diagnosed immediately after hospitalization and all of their contacts are identified and vaccinated without delays. In the absence of rapid diagnostic tests for NiV, delays in case identification would be expected, also potentially only a proportion of contacts may be identified and enrolled in the study. This however only means that the estimated trial durations (already exceeding 100 years) would be even longer when taking these limitations into account.

Improving case identification can help to reduce study durations, for example through enhanced surveillance for NiV cases (increasing the geographic area of surveillance and the number of hospitals or healthcare facilities in affected areas). Such studies could focus on groups at high-risk for zoonotic infections such as individuals collecting or consuming palm sap [32]. Healthcare workers may be an alternative high risk group for NiV infection; however they are currently only rarely infected in the setting of Bangladesh as family members act as main caregivers even for hospitalized NiV patients [6,29]. It should also be considered that based on a modelling study only about 50% of zoonotic cases are currently missed [20]. Even with perfect surveillance with all cases detected, the ring vaccination and cluster-randomized trials would remain unfeasible. Note that while the identification of asymptomatic/mild infections by serological investigations may be a viable option for some other pathogens, the incidence of such infections with NiV is estimated to be very rare and therefore would not provide substantial additional benefit [6].

While clinical trials are still considered as the dominant licensure pathway, a case-control study, if appropriately designed, can provide important insights on vaccine efficacy based on human data. This means however that the study will depend upon mass vaccination and require informed consent of participants and approval by local drug authorities. Mass vaccination as part of a vaccine trial has been previously done for the evaluation of an unlicensed cholera vaccine in Bangladesh [33]. An alternative pathway of licensing would be the use of the Animal Rule, where well designed animal studies are considered as sufficient to prove effectiveness of a vaccine [34]. Results from these animal studies can then be bridged to humans by the use of well-established correlates of immunity. This licensure pathway has been previously applied in the case of anthrax [35]. Guidance on licensing in the case where clinical trials are infeasible should be established by licensing authorities and standardized, as this problem not only exists for NiV but also other emerging pathogens.

## Data Availability

Not applicable

## Acknowledgements

B. N. and S. C. acknowledge the support of the Laboratory of Excellence Integrative Biology of Emerging Infectious Diseases (Grant ANR-10-LABX-62-IBEID), the National Institute of General Medical Sciences Models of Infectious Disease Agent Study Initiative, the AXA Research Fund and the INCEPTION project (PIA/ANR-16-CONV-0005).

## Conflict of Interest

M.L. has received grants from Pfizer, personal fees from Merck, personal fees from Bristol-Meyers Squibb, personal fees from Sanofi Pasteur, unrelated to the present work.

## Notes

### Competing Interest Statement

ML has received grants from Pfizer, personal fees from Merck, personal fees from Bristol-Meyers Squibb, personal fees from Sanofi Pasteur, unrelated to the present work.

### Author Declarations

This is a simulation study- no IRB was necessary.

