## Supplementary material for "Assessing the feasibility of Nipah vaccine efficacy trials based on previous outbreaks in Bangladesh"

1. **Parameter values**

**Table S1. Parameter values for simulations in baseline and sensitivity analysis.**

| **Parameter** | **Baseline scenario** | **Source** | **Sensitivity analysis** |
|---|---|---|---|
| **Zoonotic cases and interhuman transmission chains** | | | |
| Zoonotic spillover detection frequency- Bangladesh (annual average) | 0.04 per 1 million | Surveillance data 2007-2018 [1] | Increased zoonotic spillover detection [2] |
| Zoonotic spillover detection frequency- Rajbari & Fardipur districts (annual average) | 0.7 per 1 million | Surveillance data 2007-2018 [1] | Increased zoonotic spillover detection [2] |
| Zoonotic spillover detection frequency- Naogaon, Rangpur, and Gopolgonj districts (annual average) | 0.1 per 1 million | Surveillance data 2007-2018 [1] | Increased zoonotic spillover detection [2] |
| Birth rate (annual) | 8.4 to 19.5 per 1,000 | Birth rate predictions for 2010-2100 [3] | NA |
| Death rate (annual) | 5.6 to 14.8 per 1,000 | Death rate predictions predictions for 2010- 2100 [3] | NA |
| Reproduction number R | Negative binomial; mean 0.20 (overdispersion k 0.06) | Surveillance data 2007-2018 [1] | NA |
| Incubation period | Gamma; mean 10 days (SD 2 days) | Case data 2001-2014[4] | NA |
| Infectivity profile | Gamma, mean 4 days (SD 2 days) | Case data 2001-2014 | Mean 2 days (SD 2 days) |
| Hospitalization delay (mean days) | Gamma; mean 5 days (SD 2 days) | Case data 2007-2014[4] | Minimal case detection delay: Mean 1 day (SD 1 day) |
| **Vaccine characteristics** | | | |
| Vaccine efficacy | 90% | Preferred target product profile[5] | Minimal target product profile:70% |
| Ramp-up period | 10 days | Preferred target product profile [5] | Optimistic scenario: 5 days<br>Minimal target product profile: 14 days |
| Post-exposure effects | 0 days | | 1 to 5 days[6] |
| **Study design** | | | |
| Ring size | 90 | Average observed ring size for Ebola[7] | 50; 150 |

| | | | |
|---|---|---|---|
| Cluster size | 100,000 | | 10,000; 1,000,000 |
| Number controls | 10 | | 1-20 |
| Vaccination coverage | 70% | Routine immunization coverage[8] | 90% |
| Vaccination frequency | Every 5 years | | Every year; every 10 years |

### 2. Ring vaccination trials- Further details

**Figure S1. Required number of rings per trial arm.** Estimates are presented for a ring size of 90 individuals and a ramp-up period of 10 days. Mean hospitalization delay (Hosp); Post-exposure duration (Postexp); Vaccine coverage (Vcov).

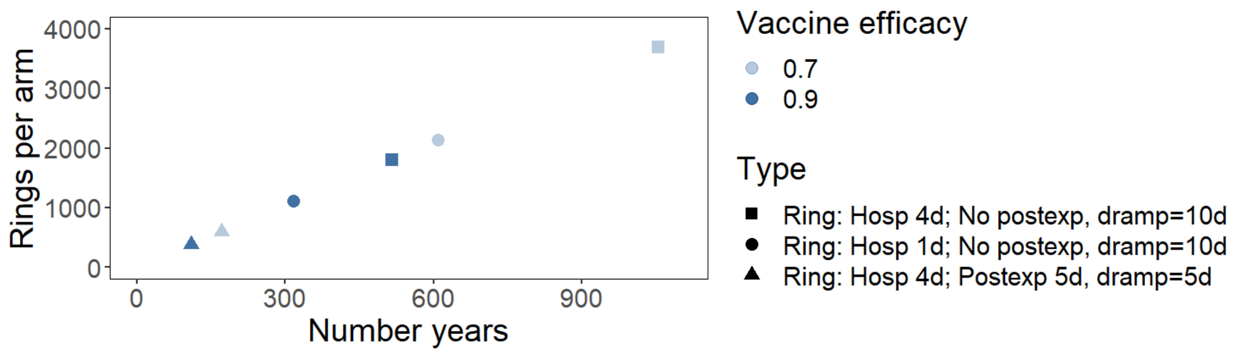


**Figure S2. Proportion of cases prevented through vaccination assuming 90% vaccine efficacy.** (A) Proportion of cases by generation prevented through vaccination in different scenarios. (B) Proportion of cases by generation resulting from 1,000 spillover simulations based on a negative binomial distribution of R=0.2 and k=0.06.

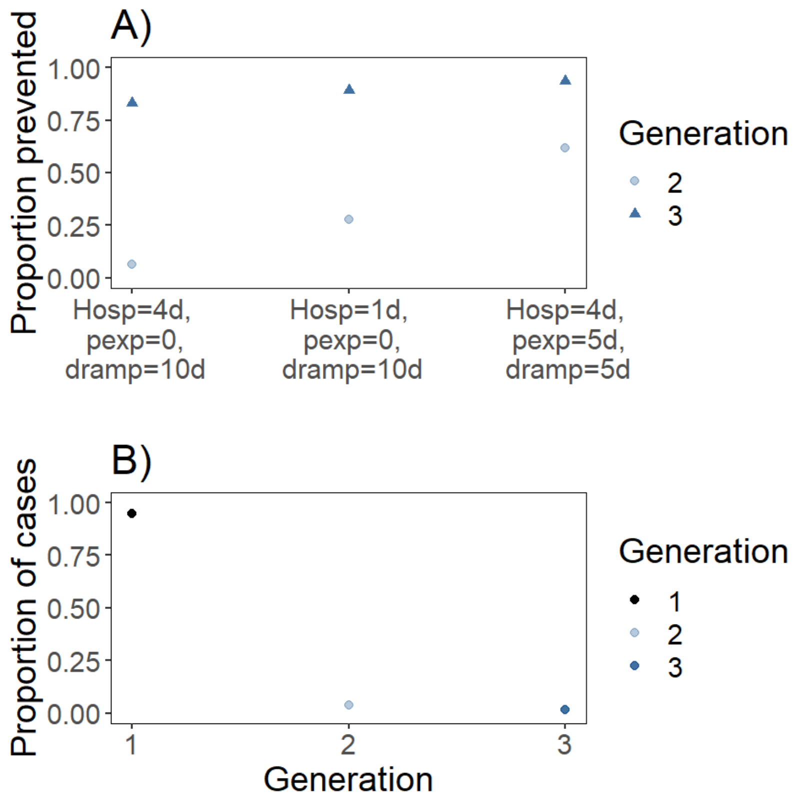

#### 3. Custer randomized trials- Further details

*Estimating the duration of the cluster-randomized controlled trial*

We simulated cluster-randomized controlled trials with 50,000 clusters per arm for 10, 20, 50, 100 and 200 years to estimate the required total population for enrolment. We then used linear regression to predict the required population size relative to the population of Faridpur and Rajbari districts for continuous trial durations up to 200 years, where the log-relative population was the outcome variable and trial duration the explanatory variable and identified the minimum duration of the trial needed to reach a log-relative population of 0.

**Figure S3. Required sample size relative to the districts' population size to conduct a cluster randomized trial within 10 years.**

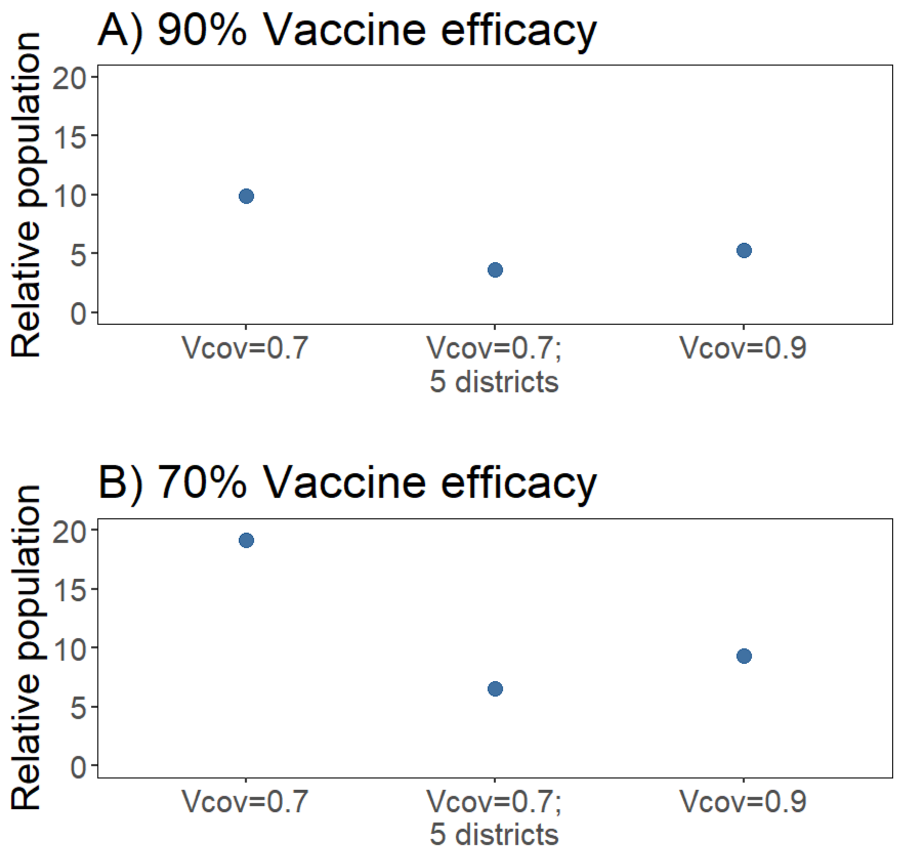

**4. Case-control studies- Further details**

**Figure S4. Number or cases required for case-control studies.**

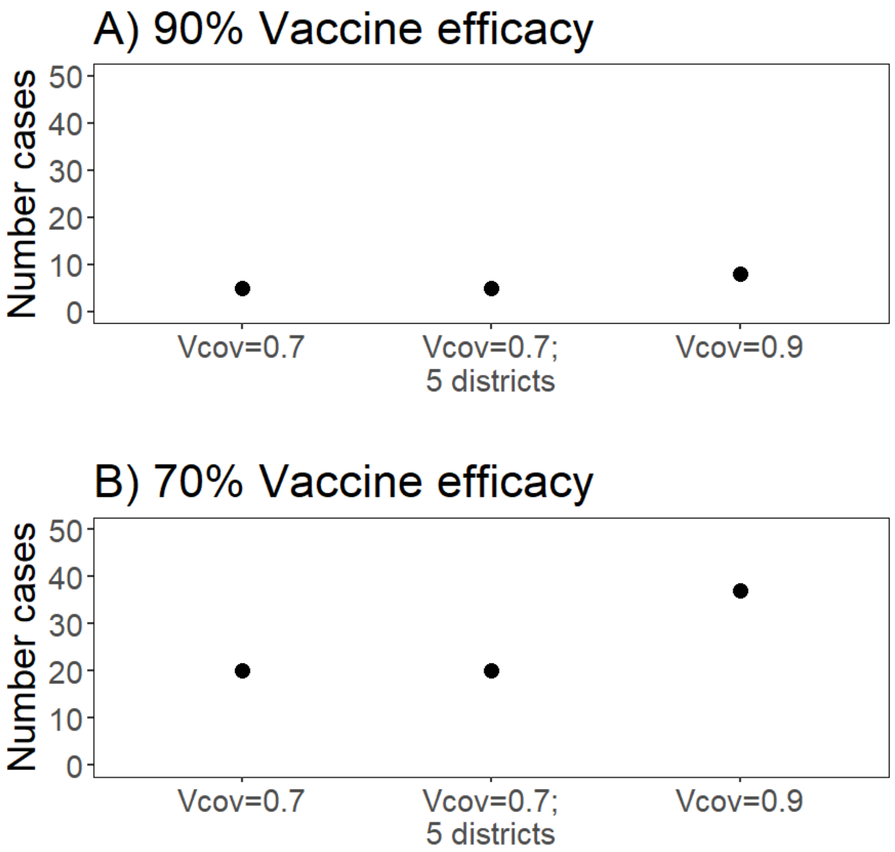

### 5. Sensitivity analysis

**Figure S5. Effect of parameter values on duration and number of doses required for ring vaccination trials.** Estimates are shown for a vaccine efficacy of 90%, a ramp-up period of 5 days, a ring size of 90 individuals and no post-exposure effects.

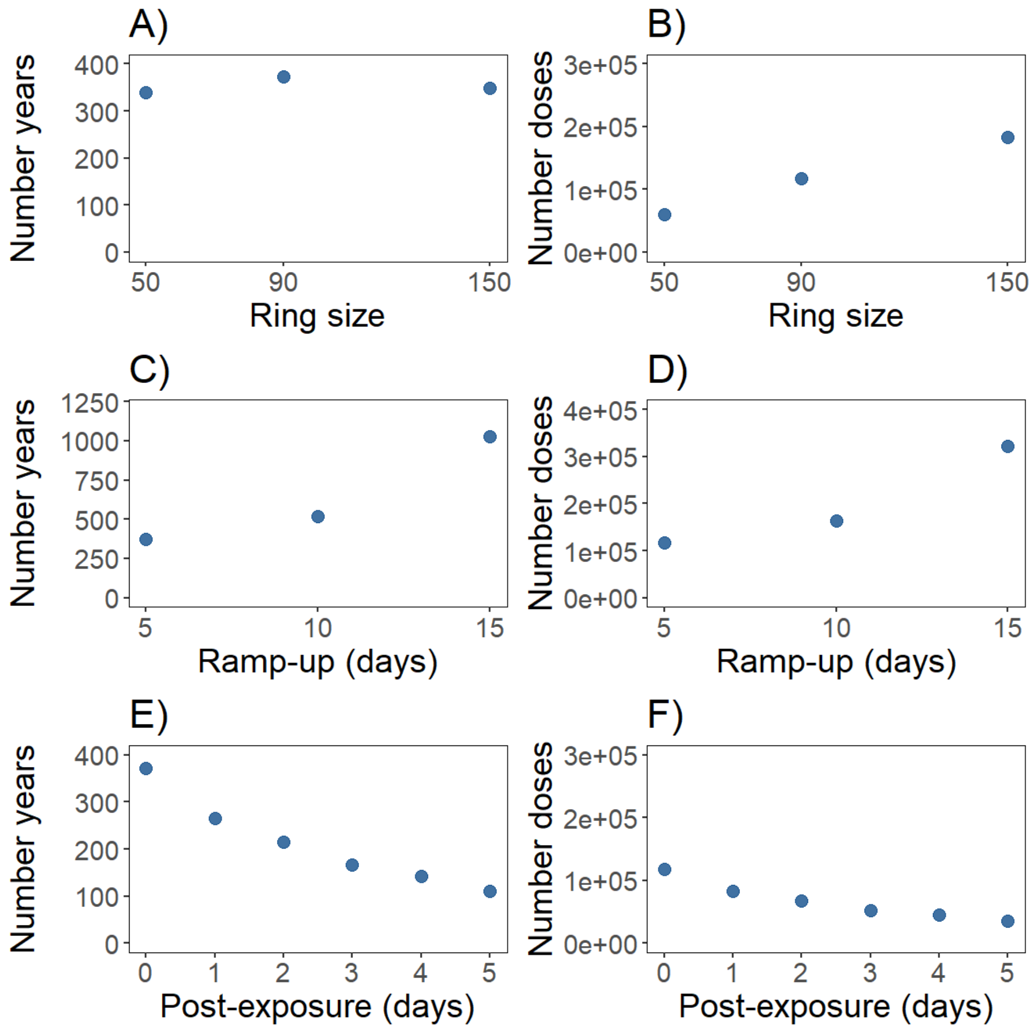

**Figure S6. Trial duration and number of doses required for a ring vaccination trial assuming an earlier peak in infectivity (infectivity profile following a gamma distribution with mean of 2 days).**

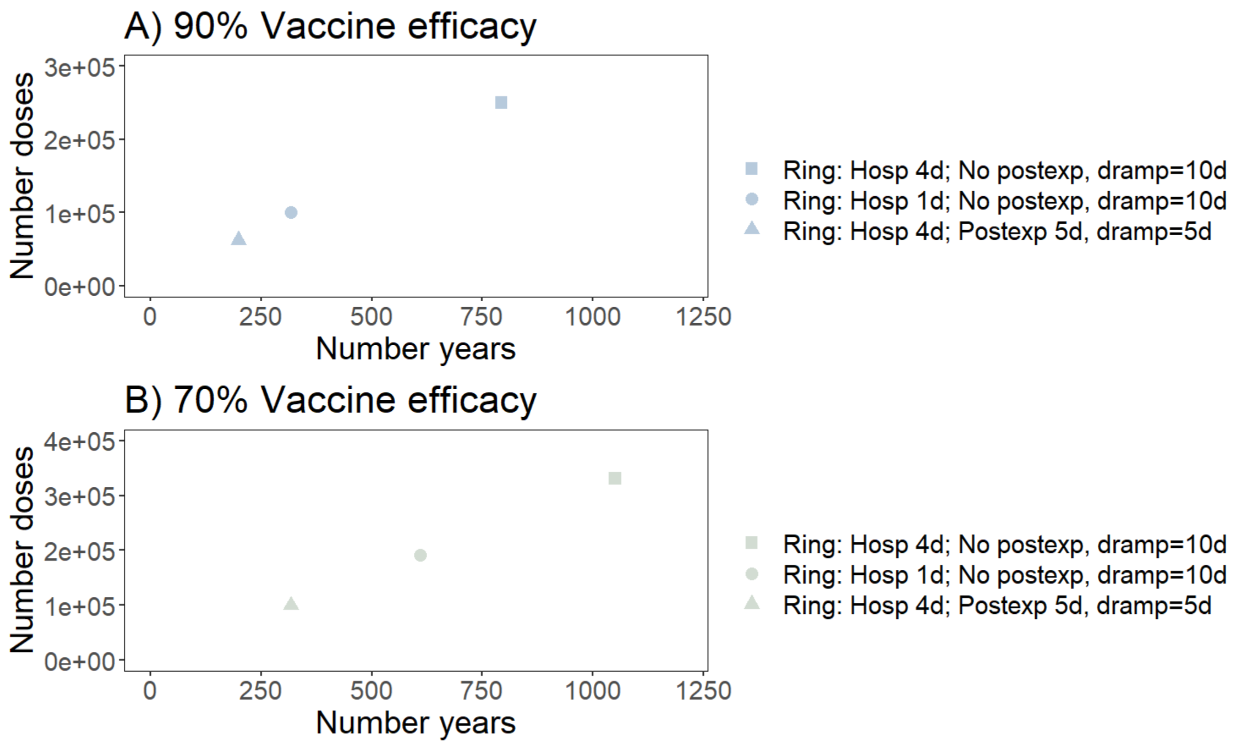

**Figure S7. Effect of parameter values on duration and number doses required for cluster randomized trials.** Estimates are shown for a vaccine efficacy of 70%, a cluster size of 100,000, a vaccination coverage of 70%, and a vaccination frequency of 5 years.

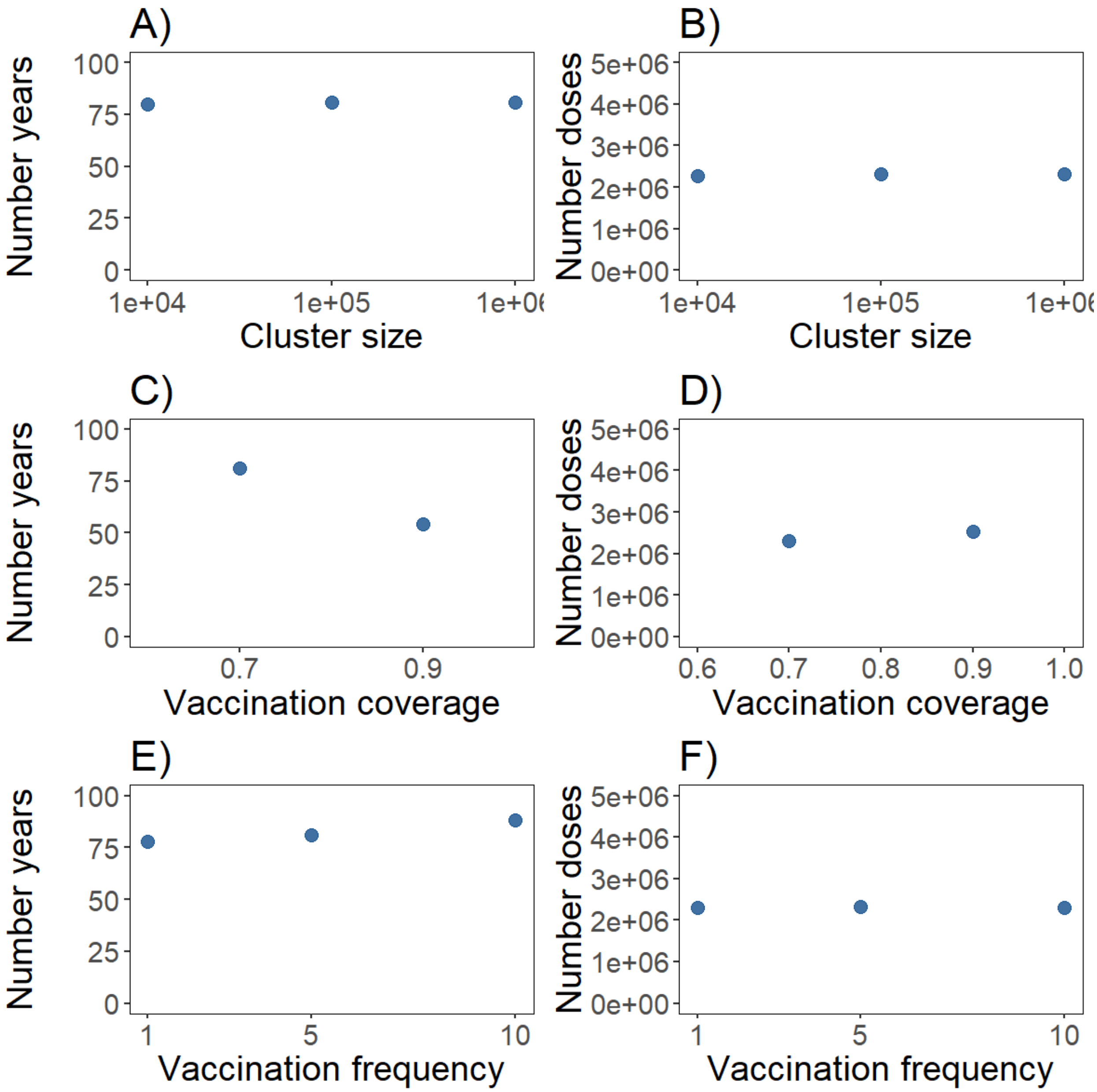

**Figure S8. Effect of parameter values on duration and number doses required for case-control studies.** Estimates are shown for a vaccine efficacy of 70%, a vaccination coverage of 70% and vaccination frequency of 5 years.

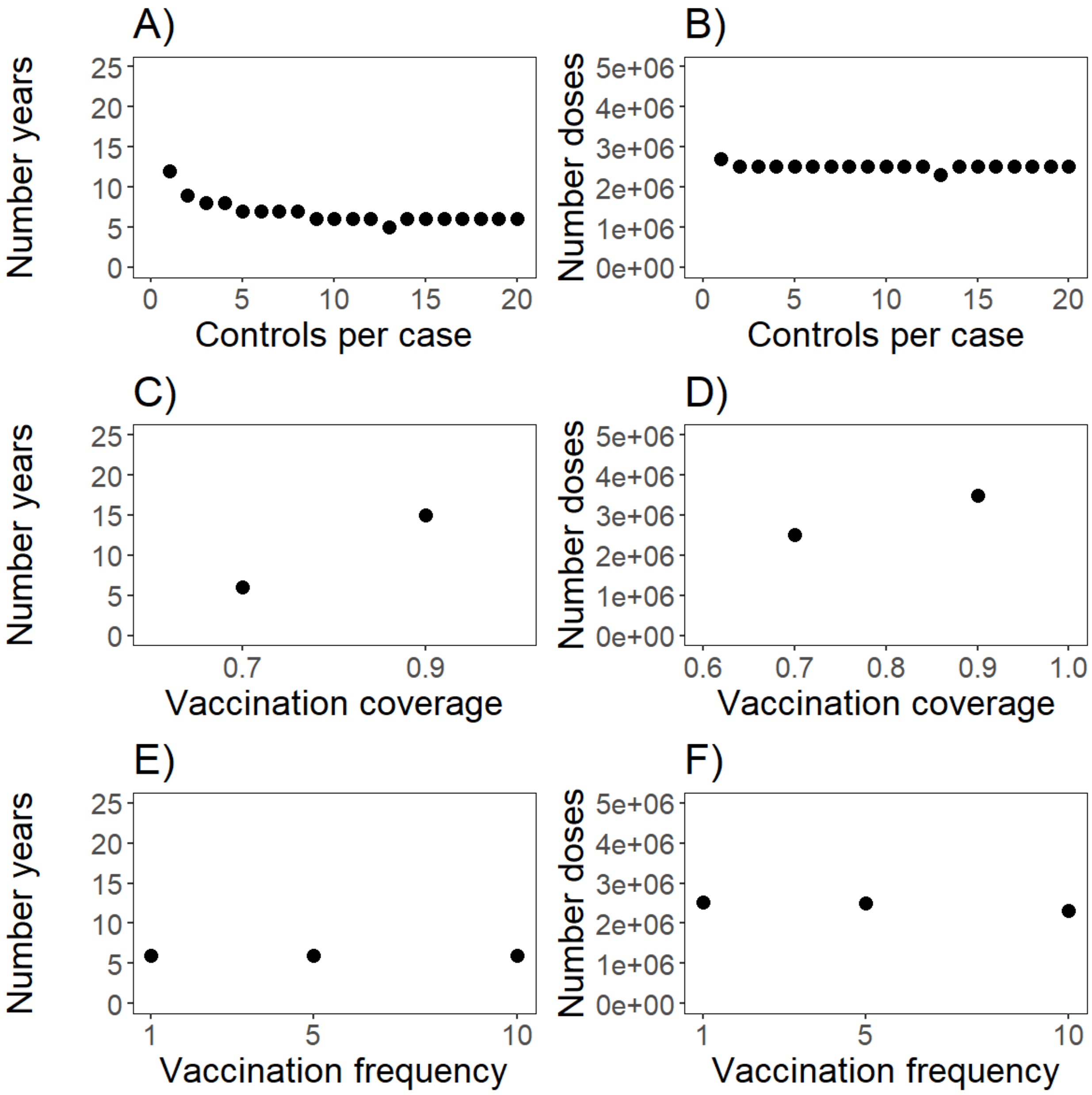
